# Aortic valve sclerosis in patients with acute myocardial infarction: a marker of increased risk of re-infarction

**DOI:** 10.1101/2023.05.15.23290018

**Authors:** Veronika A. Myasoedova, Mattia Chiesa, Nicola Cosentino, Alice Bonomi, Monica Ludergnani, Michele Bozzi, Vincenza Valerio, Donato Moschetta, Ilaria Massaiu, Valentina Mantegazza, Giancarlo Marenzi, Paolo Poggio

**Affiliations:** Centro Cardiologico Monzino, IRCCS, Milan Italy; Department of Electronics, Information and Biomedical engineering, Politecnico di Milano, Milan, Italy; Department of Clinical Sciences and Community Health, Cardiovascular Section, University of Milan, Milan, Italy

**Keywords:** acute myocardial infarction, aortic valve sclerosis, predictors, topological data analysis

## Abstract

**Background:** Patients with acute myocardial infarction (AMI) are at increased risk of recurrent cardiovascular events. Aortic valve sclerosis (AVSc), which reflects a systemic damage, may serve as a marker of risk. The aim of the present study is to better stratify sub-groups of AMI patients with specific probabilities of recurrent AMI and to evaluate the importance of AVSc in this setting.

**Methods:** We analyzed 2120 AMI patients admitted at Centro Cardiologico Monzino (2010-2019) who underwent echocardiographic evaluation for AVSc assessment. Topological data analysis (TDA) was used to stratify sub-groups of patients experiencing recurrent AMI and a random forest procedure to evaluate the importance of baseline clinical characteristics. Kaplan-Meier and Cox regression analyses were used to evaluate recurrent AMI incidence, for up to 10 years.

**Results:** TDA highlighted the presence of 8 clusters of patients with specific risks of recurrent AMI. The evaluation of time-to-event curves allowed us to combine these clusters into three super-clusters (*i*.*e*., low-, moderate-, and high-risk) and a random forest procedure showed AVSc as the most relevant variable to discriminate the three classes of risk. AVSc was detected in 1000 (47%) patients. After full adjustment, we found a significant association of AVSc with recurrent AMI (hazard ratio (HR) 1.42, 95%CI:1.03-1.97). Stratifying by age, this association was significant only for patients younger than 75 years after 5 years of follow-up (HR 1.59, 95%CI:1.03-2.46).

**Conclusions:** AVSc has been identified to be the most relevant variable to recognize patients at high risk of recurrent AMI. AVSc is frequently detected in AMI patients and is strongly associated with recurrent AMI, especially in patients younger than 75 years old. The presence of AVSc should be taken into consideration to improve the risk stratification and clinical management of AMI patients.

## INTRODUCTION

Patients with acute myocardial infarction (AMI) are at increased risk of recurrent cardiovascular events after hospital discharge, ^1^ and data from recent real-world registries reported an average 5-year all-cause mortality rate, after AMI, of ∼ 25% ^2^ and hospitalization for recurrent AMI of ∼ 30% ^3^. Therefore, it is imperative to identify the precise phenotype(s) that characterize(s) patients who will encounter adverse events after AMI and aortic valve sclerosis (AVSc) may serve as a marker of risk. AVSc is the earliest manifestation of aortic valve stenosis (AS), characterized by non-uniform thickening of the aortic leaflets without obstruction to the left ventricular outflow tract ^4^. The estimated prevalence of AVSc is around 25-30% in subjects over 65 years of age ^4,5^. The initial mechanisms involved in AVSc development, such as lipid deposition, oxidative stress, inflammation, and calcification are very similar to those of atherosclerosis ^6^. Results of our previous studies demonstrated that the atherosclerosis risk factors, such as age, hypertension, dyslipidemia, and diabetes mellitus, are associated with AVSc ^7^, while epidemiological studies suggested that AVSc is a predictor of both all-cause and cardiovascular mortality ^8,9^. Thus, it is not surprising that atherosclerotic diseases (*e*.*g*., carotid or coronary atherosclerosis) and AVSc often coexist in the same subject ^10,11^.

In this context, the application of unsupervised methodologies has been proven to be convincing in identifying (sub-)phenotypes of patients, in the cardiovascular field ^12,13^. Among them, topological data analysis (TDA) is a robust and effective unsupervised methodology, representing complex data in a low-dimensional space and preserving the intrinsic characteristics of data and the mutual relations among observations ^14^.

To better stratify sub-groups of AMI patients with specific probabilities of recurrent AMI and to evaluate the importance of AVSc in this setting, we employed the unsupervised TDA and random forest procedure. Therefore, to evaluate whether AVSc can be an independent prognostic predictor in patients with AMI or a biomarker reflecting their comorbidity burden, we analyzed more than 2100 AMI patients followed up for at least 2.5 and up to 10 years.

## METHODS

### STUDY POPULATION

This is a large prospective cohort study that consecutively recruited AMI patients, hospitalized at Centro Cardiologico Monzino IRCCS (CCM), Milan, Italy, between June 2010 and December 2019. Both ST-elevation (STEMI) and non-ST-elevation myocardial infarction (NSTEMI) patients were included in the study. Patients with significant valvular pathologies, intended as ≥ moderate valvular stenosis and/or regurgitation, major concomitant systemic conditions (*e*.*g*., malignancies), or poor echocardiographic images were excluded from the study. The study was approved by the Institutional Review Board and by the Ethical Committee of CCM (R1348/20-CCM 1418).

### TOPOLOGICAL DATA ANALYSIS (TDA)

Building topological models as networks, TDA allows complex diseases to be inspected in a continuous space, where subjects can ‘fluctuate’ over the graph, sharing more than one node of the network. In addition, TDA allows the identification of specific connected clusters (called ‘communities’) in the network, sharing similar features ^15^.

We pre-processed the original dataset for TDA as follows: 1) features with more than 5% missing values were removed; 2) binary variables where minority class exhibits a frequency lower than 2% were removed; 3) samples with more than 5 missing values were discarded; and 3) data imputation was performed on the remaining dataset, composed of 2120 samples and 35 features. Regarding the setting of TDA parameters, the first two dimensions extracted from the ‘umap’ manifold learning techniques have been chosen as lenses. The optimal configuration of TDA parameters has been obtained via a ‘grid search’ approach and assessed by the network entropy, ranging the number of ‘umap’ nearest neighbors (NN), the ‘umap’ minimal proximity between points (MP), the number of bins of the TDA map (NB), and the overlapping ratio between them (OB). The best parameters’ set was: NN=10, MP=0.3, NB=18, and OB=0.2. TDA was performed in R using the ‘PIUMA’ (https://github.com/BioinfoMonzino/PIUMA) packages. Networks have been plotted by Cytoscape ^16^.

### OBSERVATIONAL STUDY PROTOCOL

All patients underwent a two-dimensional transthoracic echocardiographic evaluation during the index event and demographic and clinical characteristics were collected during the hospitalization (*i*.*e*., baseline). The primary endpoint of the study was the re-hospitalization for recurrent AMI and was retrieved from medical records of all hospitalizations and of outpatient visits, collected during follow-up.

### ECHOCARDIOGRAPHIC EVALUATION

Two experienced cardiologists reviewed all recorded transthoracic echocardiographic images performed during index hospitalization and assessed the morphology and function of the aortic valve to evaluate the presence of AVSc, expressed as a dichotomous variable (yes or no). The AVSc was identified according to criteria described by Gharacholou *et al*. ^4^, irregular, non-uniform thickening of portions of the aortic valve leaflets or commissures, or both; thickened portions of the aortic valve with an appearance suggesting calcification (*i*.*e*., bright echoes); non-restricted or minimally restricted opening of the aortic cusps; and peak continuous wave Doppler velocity across the valve < 2 m/s.

### STATISTICAL ANALYSIS

Continuous variables are presented as mean ± SD, while categorical data are reported as frequencies and percentages. Group comparisons for continuous and categorical variables were performed by Student t-test for independent samples and by chi-square (χ2) test, respectively. Kaplan-Meier analysis was used to generate time-to-event curves and the log-rank test was used to compare strata. Univariate and multivariate Cox regressions and the related hazard ratios (HR), were calculated exploiting the ‘survival’ (v. 3.2-13) and ‘survminer’ (v. 0.4.9) R packages, while plots were generated by the ‘ggplot2’ (v. 3.3.5) R package. A p-Value < 0.05 was considered statistically significant. The data imputation and the features importance assessment were performed by ‘randomForest’ R package ^17^.

## RESULTS

### BASELINE CHARACTERISTICS

Of the 2582 patients initially examined, 2388 had good quality echocardiographic images and were not affected by valvular pathologies. After excluding patients lost to follow-up and those with more than 5 missing data, the final topological analysis included 2120 AMI patients (**Figure 1**).

**Figure 1.**
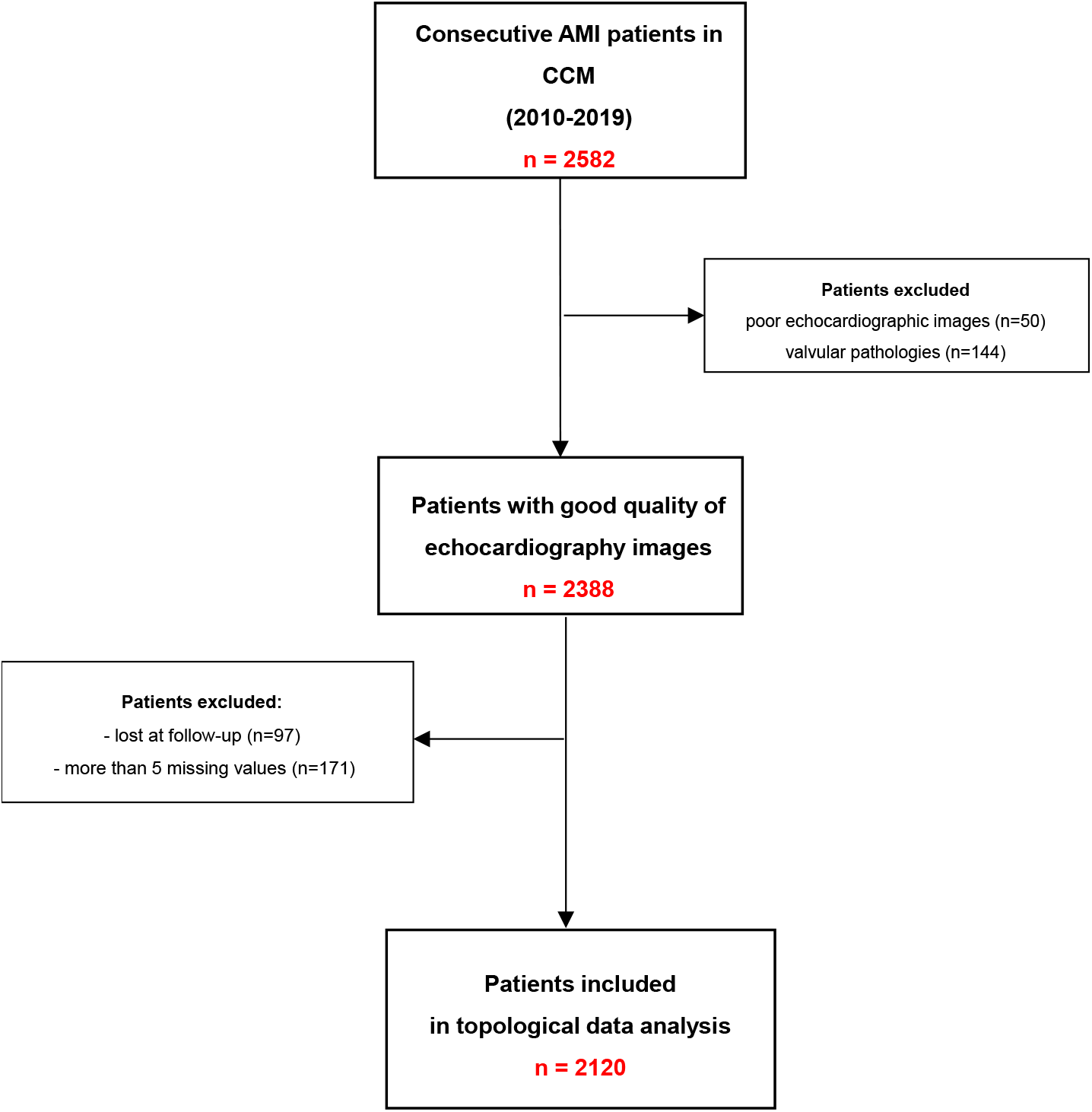
Flow diagram of the study. AMI: acute myocardial infarction; CCM: Centro Cardiologico Monzino IRCCS

### CHARACTERIZATION OF SUB-PHENOTYPES AND ASSOCIATION WITH RECURRENT AMI

Using only variables collected at baseline, we generated a network where each node represents a group of patients with similar characteristics, while edges thickness depicts the number of samples shared by nodes. After the cleaning procedure, the year-by-year analysis emphasized that recurrent AMI occurred in specific group of patients (**Supplementary Figure S1**). Since the median follow-up was two years and more than 50% of events were observed within this period, from the indexed AMI (number = 129; 56%), we focused our subsequent analysis on this particular time-frame.

TDA highlighted the presence of 8 clusters of patients who exhibited specific risks of recurrent AMI (**Figure 2A**). In particular, samples belonging to the cluster 1, 2 and 6 were labeled at higher risk of adverse event (*i*.*e*., high frequency of darker green nodes) compared to the cluster 3, 7 and 8 and, in turn, to the cluster 4 and 5, where the rate of myocardial infarction gradually decreased (majority of yellow nodes vs. green ones). This finding is also confirmed by the Kaplan-Meier event-free rate curves. The average risk of event-free rate after 2 years from discharge is close to 98%, 92%, and 83% when aggregating clusters 4 and 5 (‘Low Risk super-cluster’), clusters 3, 7 and 8 (‘Moderate Risk super-cluster’), and cluster 1, 2 and 6 (‘High Risk super-cluster’), respectively (**Figure 2B**). Consequently, the High Risk super-cluster shows an average HR of 5 (95% CI: 2.2-11.5), while the Moderate Risk super-cluster has an average HR of 2.5 (95% CI: 1.1-5.9) when compared to Low Risk super-cluster, used as reference (**Figure 2C**).

**Figure 2.**
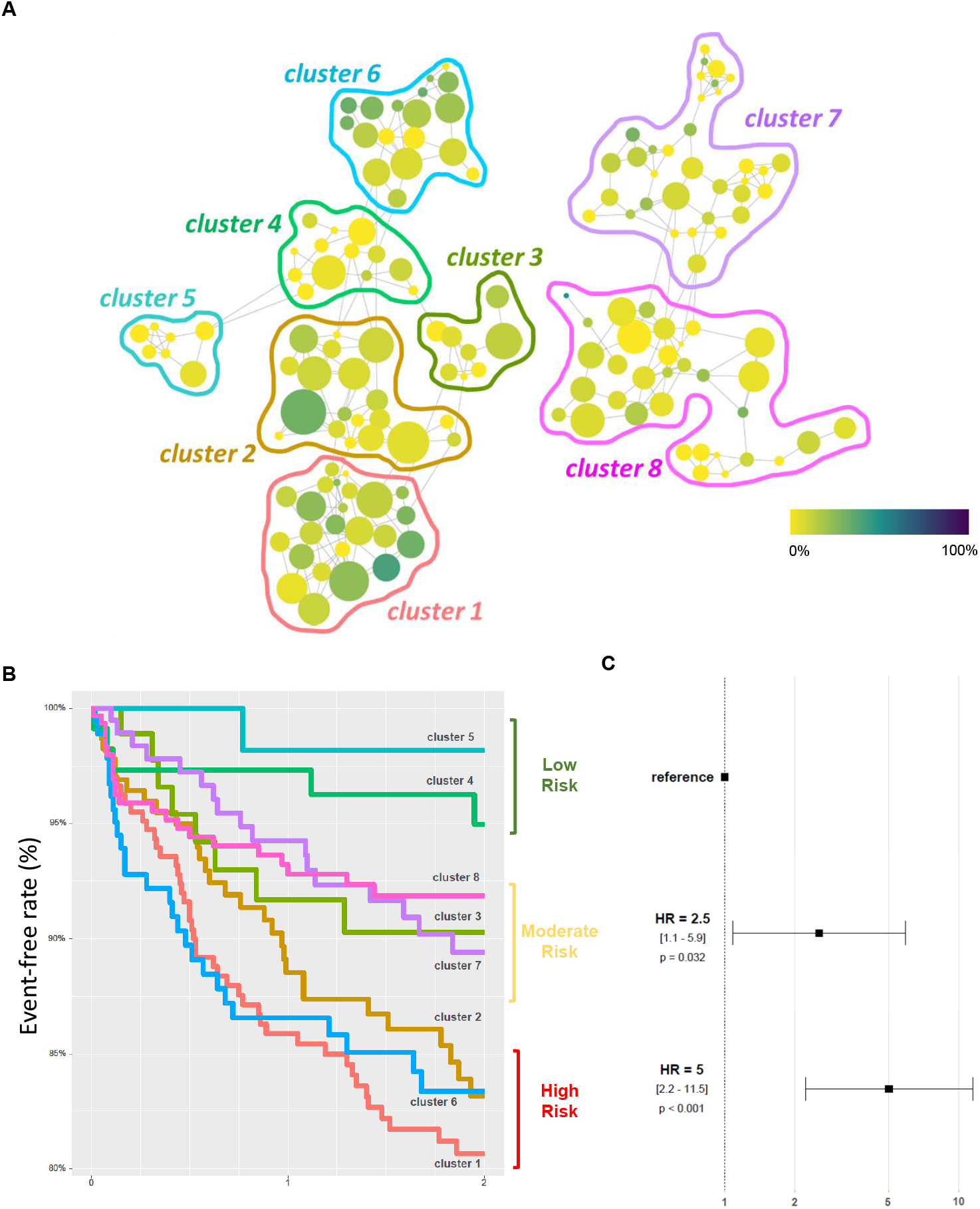
AMI patients and sub-phenotypes identification. (**A**) TDA results shown as network. Each node represents a group of patients with similar clinical characteristics, while edges thickness depicts the Jaccard’s index (*i*.*e*., the number of samples shared by nodes). The node color represents the average risk of recurrent AMI after 2 years from discharge between samples belonging to a specific node from 0 (yellow nodes) to 1 (purple nodes). Each identified cluster has been highlighted with different colors. In this network, nodes with less than 3 neighbors with Jaccard’s index = 1 has been removed to reduce noise. (**B**) Kaplan-Meier curves show the event-free survival for each cluster at 2 years follow-up. Three ‘super-cluster’ has been defined grouping cluster with similar event-free survival (*i*.*e*., ‘High Risk’, ‘Moderate Risk’ and ‘Low Risk’). (**C**) Hazard ratio from Cox regression at 2 years follow-up for ‘High Risk’ and ‘Moderate Risk’ compared to ‘Low Risk’ super-cluster used as reference.

The occurrence of separated clusters belonging to the same class of risk (*i*.*e*., super-clusters) suggests that different combinations of clinical features could lead to the same probability of adverse event. Indeed, we found that the patients belonging to the High Risk super-cluster are most frequently men with hypertension and hypercholesterolemia. However, clusters 1 and 2 shows a higher number of patients with previous AMI, NSTEMI at indexed event and AVSc presence, while in clusters 2 and 6 there are patients with diabetes mellitus, a lower level of high-density lipoprotein (HDL) and a higher body mass index (BMI). Moreover, patients belonging to the Moderate Risk super-cluster share a higher level of high-sensitive C reactive protein (hsCRP, except cluster 7; **Figure 3A**). Of note, AVSc was highly prevalent in cluster 1, 2, 3, and 7 referring to Moderate and High Risk super-clusters.

**Figure 3.**
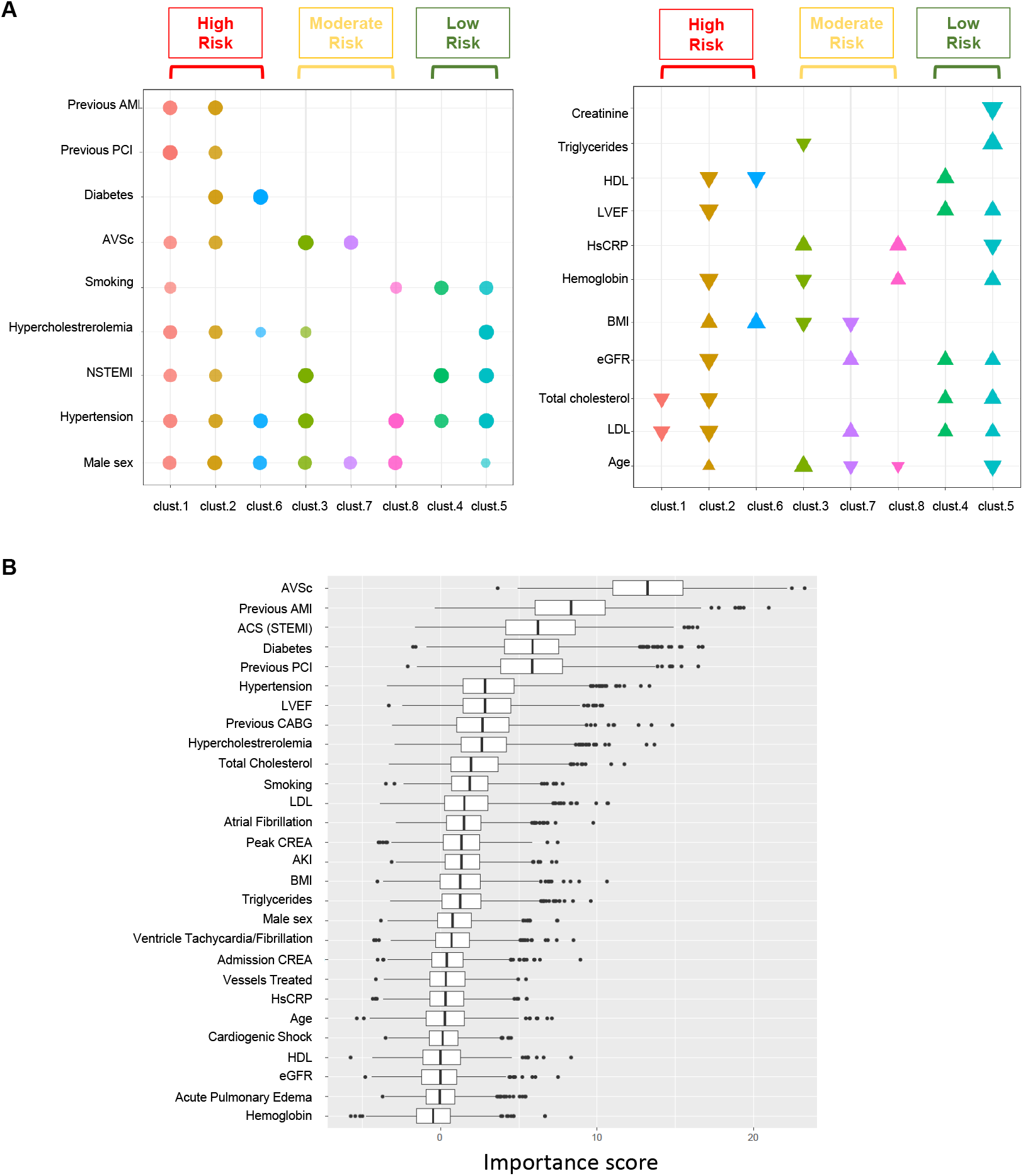
AMI patients and sub-phenotypes characterization. (**A**) Bubble plot where the circle size depicts the frequency of samples exhibiting a specific feature in each cluster. Circles with a frequency lower than 0.6 have been removed. Bubble plot where the tringle size depicts the absolute value, scaled over all clusters and the tringle directions represent the sign for each continuous variable. Tringles with an absolute scaled value greater than 0.65 were removed. (**B**) Features Importance. Box plot showing the importance score in terms of mean decrease in accuracy, when trying to classify three super-clusters: ‘Low Risk’, ‘Moderate risk’, and ‘High Risk’. The score has been generated by a Random Forest procedure repeated 1000 times.

Finally, we assessed which variables were the most relevant to discriminate the three classes of risk, implementing a random forest procedure 1000 times, and then, calculating the importance score of each feature. The resulting importance scores (IS) represent the percentage accuracy loss in risk prediction for the specific risk factors. As summarized in **Figure 3B**, AVSc reached the highest average importance score (aIS = 13.2) compared with well-known risk factors for recurrent AMI, such as previous AMI (aIS = 8.5), diabetes mellitus (aIS = 6.1), arterial hypertension (aIS = 2.7), and left ventricular ejection fraction (LVEF; aIS = 2.6), suggesting that without AVSc we will lose ∼ 13% of accuracy in predicting the risk of recurrent AMI.

### AORTIC VALVE SCLEROSIS AND RECURRENT AMI

We assessed the association between long-term recurrent AMI and the presence of AVSc in 2120 AMI patients. AVSc was presented in 1000 (47%) of the AMI patients (**Table 1**). As expected, patients with AVSc were older and had more frequent cardiovascular risk factors as well as previous AMI. Moreover, patients with AVSc had significantly lower LVEF and estimated glomerular filtration rate (eGFR) compared to patients without AVSc. Finally, AVSc patients had a more complicated in-hospital clinical course.

**Table 1.** Baseline clinical characteristics and in-hospital outcomes of study patients with and without AVSc.

| Variable | No-AVSc<br>n=1120 | AVSc<br>n=1000 | p-Value |
| --- | --- | --- | --- |
| Age, years | 61.7±11.5 | 72.2±10.9 | <0.001 |
| Male sex, n (%) | 877 (78) | 712 (71) | <0.001 |
| Body mass index, kg/m<sup>2</sup> | 26.8±4.4 | 26.3±4.1 | 0.050 |
| Diabetes mellitus, n (%) | 197 (18) | 250 (25) | <0.001 |
| Hypertension, n (%) | 620 (55) | 706 (71) | <0.001 |
| Smoking, n (%) | 666 (60) | 475 (48) | <0.001 |
| Dyslipidemia, n (%) | 495 (44) | 501 (50) | 0.007 |
| Prior AMI, n (%) | 188 (17) | 294 (29) | <0.001 |
| Peripheral artery disease, n (%) | 23 (2.1) | 36 (3.6) | 0.043 |
| Prior CABG, n (%) | 56 (5) | 153 (15) | <0.001 |
| Prior PCI, n (%) | 208 (19) | 294 (29) | <0.001 |
| STEMI, n (%) | 608 (54) | 508 (50) | 0.119 |
| Time-to-presentation, hours | 10.5±23.2 | 11.2±22.1 | 0.501 |
| Left ventricular ejection fraction, (%) | 51±11 | 49±12 | <0.001 |
| PCI during index hospitalization, n (%) | 920 (82) | 727 (73) | <0.001 |
| 3-vessel disease, n (%) | 26 (2.3) | 26 (2.6) | 0.679 |
| **Admission laboratory values** |  |  |  |
| Total Cholesterol, mg/dL | 182±44 | 171±43 | <0.001 |
| LDL, mg/dL | 115±39 | 105±39 | <0.001 |
| HDL, mg/dL | 42±13 | 44±14 | 0.009 |
| Triglycerides, mg/dL | 126±73 | 115±66 | <0.001 |
| Blood glucose, mg/dL | 126±46 | 131±54 | 0.307 |
| HbA1c | 6.1±1.4 | 6.1±1.2 | 0.075 |
| Creatinine, mg/dL | 1.0±1.4 | 1.1±0.6 | <0.001 |
| eGFR, mL/min/1.73 m<sup>2</sup> | 82±23 | 73±25 | <0.001 |
| Hemoglobin, g/dL | 14.1±1.7 | 13.5±1.9 | <0.001 |
| High-sensitivity C-reactive protein, mg/dL | 15.9±43.9 | 18.6±43.1 | 0.004 |
| **In-hospital complications** |  |  |  |
| Cardiogenic shock, n (%) | 52 (4.6) | 62 (6.2) | 0.136 |
| Mechanical ventilation, n (%) | 33 (2.9) | 47 (4.7) | 0.045 |
| Blood transfusion, n (%) | 21 (1.9) | 33 (3.3) | 0.052 |
| Ventricular fibrillation/ventricular tachycardia, n (%) | 86 (7.7) | 66 (6.6) | 0.381 |
| Atrial fibrillation, n (%) | 80 (7.1) | 165 (16.5) | <0.001 |
| Acute pulmonary edema, n (%) | 60 (5.4) | 121 (12.1) | <0.001 |
| Acute kidney injury, n (%) | 91 (8.1) | 121 (12.1) | 0.003 |
| High-degree atrio-ventricular block, n (%) | 31 (2.8) | 44 (4.4) | 0.056 |
| CCU length of stay, days | 4.0±2.6 | 4.1±2.3 | 0.021 |
| **Discharge therapy** |  |  |  |
| Statins, n (%) | 1051 (94) | 1035 (91) | 0.004 |
| Dual antiplatelet therapy, n (%) | 1046 (93) | 907 (91) | 0.073 |
| Beta-blockers, n (%) | 900 (80) | 769 (77) | 0.118 |
| ACE/ARB, n (%) | 717 (64) | 666 (67) | 0.122 |

ACE/ARB = angiotensin-converting enzyme inhibitors/angiotensin receptor blockers; CABG = coronary artery bypass graft; CCU = coronary care unit; eGFR = estimated glomerular filtration rate; AMI = acute myocardial infarction; PCI = percutaneous coronary intervention; STEMI = ST-elevation myocardial infarction.

We found that patients with AVSc had an increased risk of recurrent AMI after 2 years of follow up (number of events = 129) in the unadjusted and age and sex adjusted models (HR 1.79, 95% CI: 1.26-2.54, p = 0.001 and HR 1.51, 95% CI: 1.03-2.22, p = 0.037, respectively) but not in the full adjusted one (HR 1.46, 95% CI: 0.93-2.30, p = 0.100; **Figure 4**). The full adjusted model comprise age, sex, BMI, STEMI, atrial fibrillation, acute pulmonary edema, diabetes mellitus, hypertension, smoking, dyslipidemia, previous AMI, chronic kidney disease, peripheral artery disease, LVEF, treated vessels, creatinine, acute kidney injury, hemoglobin, hsCRP, statins, and dual antiplatelet therapy.

**Figure 4.**
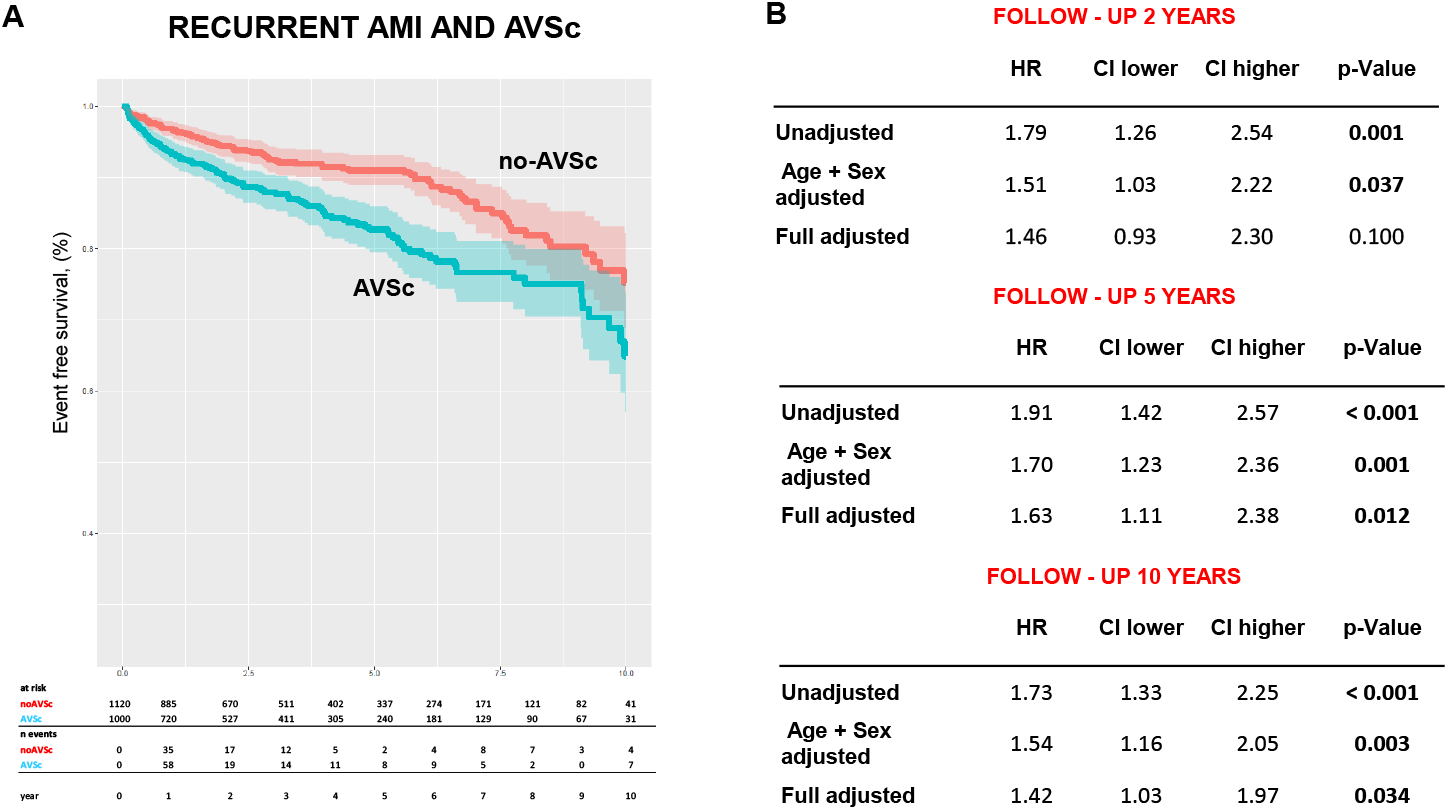
Recurrent AMI and AVSc. (**A)** The Kaplan-Meier curves showing the incidence of re-infarction in AMI patients with and without AVSc. (**B**) The plots show the strength of the associations between re-infarction and AVSc at 2, 5 and 10-year follow-up (hazard ratio -HR) and their distribution.

Nevertheless, at 5 and 10 years of follow-up AVSc was significantly associated with recurrent AMI even in the full adjusted models (HR 1.63, 95% CI: 1.11-2.38, p = 0.012 and HR 1.42, 95% CI: 1.03-1.97, p = 0.034 respectively).

Remarkably, AVSc was significantly associated with recurrent AMI only in patients younger than 75 years of age (**Figure 5**). Specifically, after 2 years of follow-up, the fully adjusted model revealed a trend in the association between AVSc and recurrent AMI (HR 1.68, 95% CI: 0.99-2.85, p = 0.053), which become significant at a 5-year follow-up (HR 1.59, 95% CI: 1.03-2.46, p = 0.036). However, probably due to advancing age, at 10 years of follow-up, the association between AVSc and recurrent AMI attenuates (HR 1.39, 95% CI: 0.96-2.02, p = 0.083).

**Figure 5.**
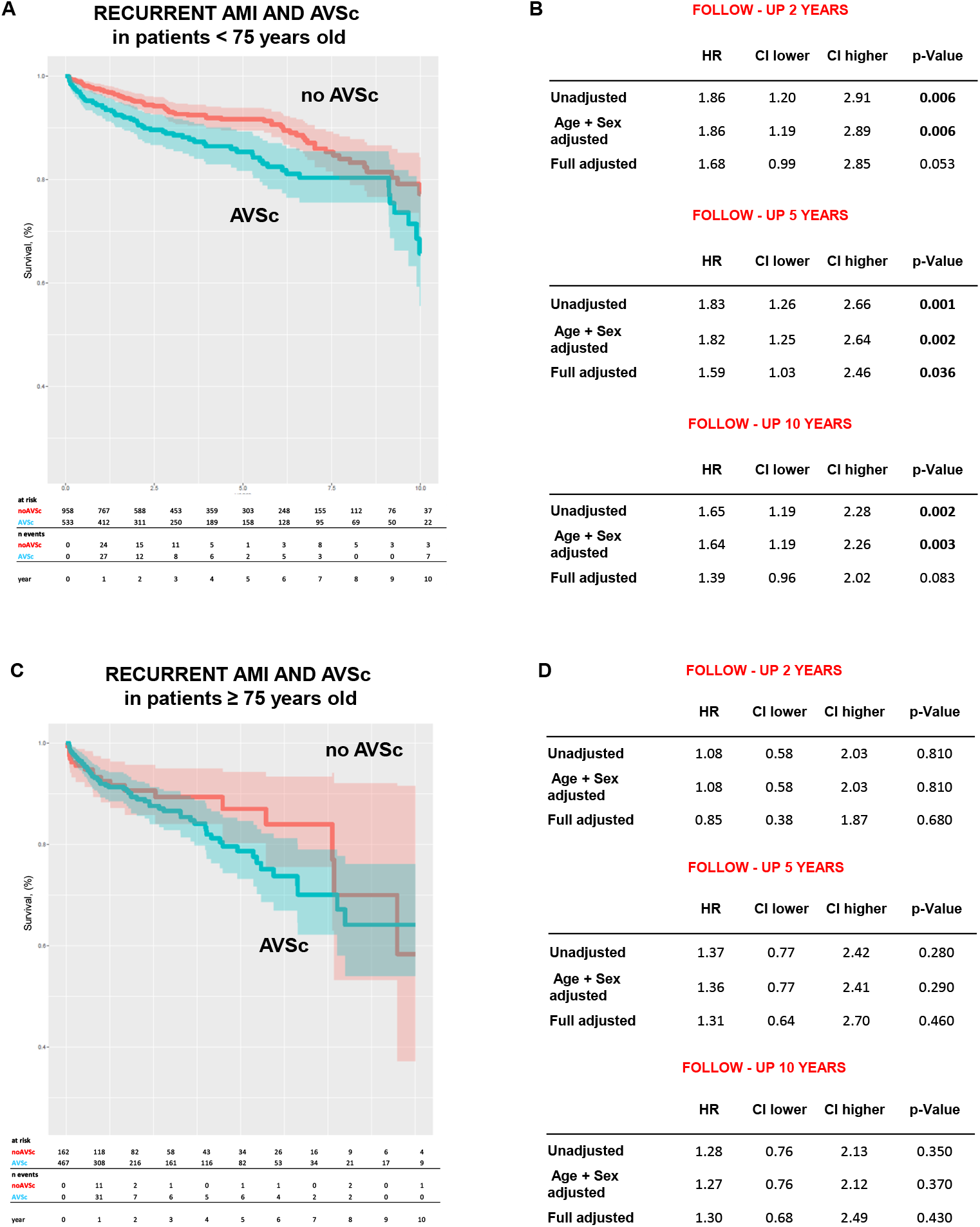
Recurrent AMI and AVSc in patients younger and older than 75 years old. (**A**) The Kaplan-Meier curves showing the incidence of re-infarction of AMI patients younger than 75 years of age with and without AVSc. (**B**) The plots show the strength of the associations between re-infarction and AVSc at 2, 5 and 10-year follow-up (hazard ratio -HR) and their distribution in AMI patients younger than 75 years of age. (**C**) The Kaplan-Meier curves showing the incidence of re-infarction of AMI patients older than 75 years of age with and without AVSc. (**D**) The plots show the strength of the associations between re-infarction and AVSc at 2, 5 and 10-year follow-up (hazard ratio -HR) and their distribution in AMI patients older than 75 years of age.

## DISCUSSION

We deeply analyze a large AMI patient cohort by applying TDA and three classes of recurrent AMI risk were identified with this unsupervised methodology. Patients in the High risk class were mostly male and had a higher prevalence of classic risk factors such as hypertension, hypercholesterolemia, diabetes, and previous AMI, whereas Low risk patients had fewer comorbidities. Interestingly, we found that AVSc was an important player in risk prediction of recurrent AMI in both Moderate and High risk groups. In addition, when we ranked these three risk classes using the importance score of the most relevant variables associated with recurrent AMI, AVSc was shown to have the highest value at 2-year follow-up. In addition, the AVSc presence, at baseline, led to a significant increase risk of recurrent AMI after 5 and 10 years of follow-up. However, the predictive ability of AVSc was preserved only in patients younger than 75 years of age.

Previously, the results of a meta-analysis on more than 30 studies, which included 10537 patients with AVSc and 25005 controls, showed that the presence of AVSc is associated with coronary artery disease, stroke, and cardiovascular mortality ^8^. Indeed, the prevalence of AVSc in patients with CAD is about 45% and even higher (> 60%) in patients with carotid atherosclerosis and after coronary or carotid revascularization these patients had increased overall mortality compared with patients without AVSc ^10,11^. Moreover, in patients who underwent coronary artery bypass graft (CABG), the 90-day survival was significantly lower in the AVSc group and the addition of AVSc to the EuroSCORE II improved the stratification of these high-risk patients ^9^.

Patients with AMI remain at very high risk of experiencing recurrent cardiovascular events after discharge and is very important to better identify and stratify their risk. To this goal, Steen *et al*. ^1^ conducted an elegant study of event rates and risk factors for recurrent cardiovascular events using several approaches in a large and well-characterized population of more than 240000 AMI patients. The authors suggested that the 5-year incidence of non-fatal AMI, non-fatal ischemic stroke, or cardiovascular death was 33% and the risk of recurrence of these events was higher immediately after discharge. Accordingly, even if our cohort was considerably smaller, we observed that the majority of recurrent events occurred within the first years after discharge.

Regarding the association between AVSc and recurrent AMI, Dursan *et al*. ^18^ reported, in a small cohort, that patients with previous AMI were more likely to have AVSc at the indexed AMI event. We directly associated AVSc presence and AMI recurrency in patients younger than 75 years of age, following these patients up to 10 years after the indexed event. Therefore, to improve the life-expectancy of patients after an AMI, new markers associated with recurrent events such as AVSc should be considered and included in overall clinical management. However, no association between AVSc and recurrent AMI in patients older than 75 years of age was noted but we should take into consideration that advanced age is one of the most negative prognostic factors for mortality following AMI ^19,20^ and thus, age could mask the prognostic effect of AVSc.

The incorporation of novel biomarkers for patient risk stratification is crucial in modern medical practice. Presently, routine echocardiography is performed on all patients admitted to hospitals with AMI, providing a simple and effective means of assessing AVSc. The identification of AVSc through echocardiography has the potential to enable early identification of patients at high risk of recurrent cardiovascular events. However, prior to the implementation of AVSc in patient management decisions, dedicated clinical trials are necessary. These trials should examine the specific pharmacological treatment of AMI patients with AVSc and compare their outcomes to those of non-AVSc patients with AMI.

## Data Availability

The data underlying this article will be shared upon reasonable request to the corresponding author.

## Limitations

Several limitations of the study should be acknowledged. First, this is a single-center study, which may limit the generalizability of the findings. However, the large number of participants enrolled in the study helps to reduce potential bias. Second, the models used in the study do not account for changes in baseline treatment that may have occurred during the lengthy follow-up period. Third, the assessment of AVSc was conducted using a dichotomous variable, as the most commonly used definition is still too broad. More accurate and unbiased methods for quantifying AVSc are needed. Fourth, different coronary stents and antithrombotic agents were used. Yet, this corresponds to a “real-world” scenario where patients are treated with different antiplatelet drugs and stents according to clinical setting, operator choice, and drug/device availability. Lastly, no information was available regarding patients’ adherence to treatment during follow-up. In particular, patients were considered to be on dual antiplatelet therapy according to the discharge treatment.

## Conclusion

Our study provides evidence that AVSc is a crucial variable for identifying AMI patients at moderate-to-high risk of recurrent AMI. AVSc is frequently observed in AMI patients and is strongly associated with re-infarction, especially in those younger than 75 years old. Therefore, our findings suggest that the presence of AVSc should be taken into account when assessing the risk of recurrent AMI and managing AMI patients. Incorporating AVSc into risk stratification models may improve the accuracy of predicting recurrent AMI and could enhance personalized treatment decisions for AMI patients.

## Findings

This work was supported by the Italian Ministry of Health funds (Ricerca Finalizzata: GR-2019-12370560 and GR-2018-12366423) and Ricerca Corrente. P.P. is supported by Fondazione Gigi e Pupa Ferrari ONLUS (FPF-14).

---

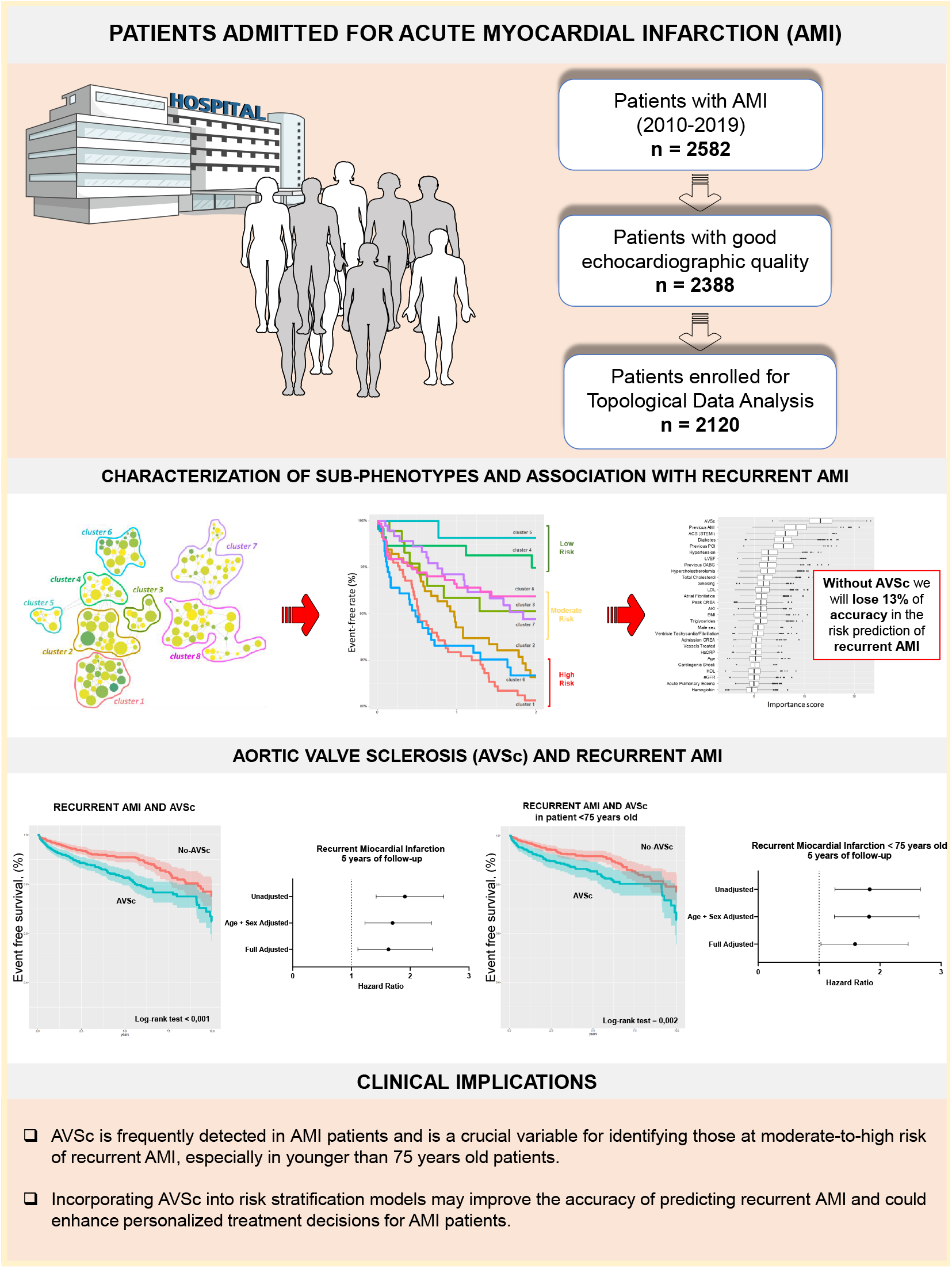

